# The COVID-19 pandemic has increased health inequalities by altering patterns of engagement with a mass participation event after resumption post-lockdown

**DOI:** 10.1101/2023.08.30.23294836

**Authors:** Andre S. Gilburn

## Abstract

The global epidemic in physical inactivity has been exacerbated by lockdowns with concerning health inequalities accentuated as a result. Studies are needed of post-lockdown levels of activity to determine if there are lasting impacts beyond lockdown. This study uses parkrun in Scotland as a model system to address this urgent question. Generalised linear mixed models were used to identify patterns in the attendance, gender, age and performance of 500,000+ parkrun participations in Scotland for the year before the pandemic and the first year after lockdown. Total weekly attendance in Scotland had been increasing year-on-year but fell by 13% from 7,649 (95% CI 7,275 to 8.024) in the final-year pre-lockdown to 6,612 (95% CI 6,260 to 6,963) in the first-year post-lockdown. Attendance drops were disproportionately larger at events with larger attendances prior to lockdown. There was a disproportionate loss of female participants in more deprived areas. The gender gap in participation had been narrowing before lockdown but widened from 54.02% (95% CI 53.85 to 54.19%) in the final-year pre-lockdown to 56.29% male (95% CI 56.09 to 56.49%) in the first-year post-lockdown. The age of participants increased from 44.94 (95% CI 44.90 to 44.98) before lockdown to 46.58 (95% CI 46.53 to 46.64) after lockdown. The age gap between the genders widened after lockdown particularly at larger events and in more deprived areas. Performance was declining before lockdown but increased from 55.02% (95% CI 54.96 to 55.06%) to 55.93% (95% CI 55.88 to 55.97%) after lockdown. The pandemic has reduced attendance at parkrun in Scotland and the loss of participants has been non-random increasing health inequalities affecting women, the least fit and those in deprived areas. Some former participants are also now avoiding larger mass participation events. Investment and management strategies will be needed to address these worsening inequalities.

## Introduction

Variation in levels of physical activity have been found to be driving health inequalities highlighting the importance of tracking levels of activity and identifying correlates of activity among populations [1–3]. The COVID-19 pandemic had a major impact on levels of physical activity [4–15]. Among the identified impacts of lockdown is an avoidance of larger events found in runners in Greece [4]. A study tracking physical activity levels in the UK found it declined 37% during lockdown, although the impacts varied with age, with older individuals more likely to maintain their pre-lockdown levels of activity than younger adults [5]. The impacts of the pandemic have also been found to vary with socio-economic background with those in deprived areas finding it hardest to access places to exercise during lockdowns [6–8]. By contrast, a study of levels of physical activity in Kenya found that people from higher social classes were more likely to have reduced their activity during lockdown and this was particularly the case for females [14]. A study of factors associated with physical activity during lockdown in Australia noted the importance of connections to an exercise group for older adults to maintain or even increase levels of activity [15]. The impacts of the pandemic on levels of physical activity exacerbate the pre-existing problem with 1.4 billion adults estimated to have been doing too little physical exercise prior to the pandemic [16]. This had resulted in the promotion of wider participation in sport becoming a global priority [17]. The negative impact of the pandemic on levels of physical fitness has only served to increase this global priority. This leaves a key question. If lockdown impacted the demographics of those engaging in physical exercise do those impacts extend into the post-lockdown era [10]? Large-scale studies investigating changes in the patterns of exercising are urgently needed to address this question [12].

One very large dataset on activity levels is the parkrun results database. parkrun provide free weekly 5km events at nearly 2,000 locations generating data for close to 100,000 events annually. There is huge potential for using parkrun as a model system for investigating changes in the demographics of those engaging in physical activity and identifying correlates of activity. Several studies have investigated the parkrun results database [18–21] and some studies have identified correlates of participation on a very large scale [22–28]. Few studies though have investigated changes in the demographics of parkrun participants over time [22,29].

A survey of the behaviour of previous parkrun participants during lockdown found reduced activity levels [29]. Increased inactivity during lockdown was particularly evident in younger adults, people in deprived areas and the least active [29]. Consequently, the changing patterns in activity among parkrun participants during lockdown are consistent with those identified in other studies [4–8] revealing the potential value of using parkrun as a model system. As more than 50,000 parkrun events have taken place in the UK alone since lockdown ended a huge database has been generated on post-lockdown activity levels at a mass participation event that not only has a key aim of encouraging the least active to engage in physical activity but is also actively involved with social prescribing [30–32]. This allows a unique opportunity to investigate what lasting impacts lockdowns have had on levels and correlates of physical activity after they have been lifted and how lockdown might have altered the demographic of parkrun participants. This is particularly important as lockdowns are likely to have increased equalities in social prescription [33].

A previous study of all parkrun events in Scotland from 2004-2019 revealed various changing correlates of participation with a narrowing gender ratio, reducing performance scores and an increasing mean age of participants [22]. Another study of factors affecting the likelihood of returning to parkrun for new participants in Scotland found that they are less likely to return after attending larger events [28]. The pandemic could have increased risk averse behaviour and exacerbated avoidance of larger parkrun events. The aim of this study is to investigate how lockdown has impacted these changing demographics and correlates of participation at parkrun in Scotland. Scotland has a range of different parkrun types from city centre parks to remote island locations. Scottish parkrun locations also show considerable variation in the level of deprivation of the surrounding areas in which they are located. This makes Scottish parkrun events an ideal model system for investigating lasting impacts of lockdown on the levels of physical activity by comparing characteristics of participants before and after lockdown. Scottish parkrun events also vary considerably in their mean number of attendees allowing an assessment of whether some former participants are not avoiding larger parkrun events to reduce the risk of exposure to COVID-19.

## Methods

This was an analytical study of aggregated publicly available secondary data. There was no primary data collection and no active participants.

### Data sources

The primary data source was the parkrun results for events that occurred in Scotland in the year immediately prior to lockdown (16^th^ March 2019 to 14^th^ March 2020) and the year immediately after the resumption (14^th^ August 2021 to 13^th^ August 2022). Data from 44 events were included. These were all the events that were in existence one year prior to the start of the pandemic and which also returned after lockdown. The results for each event were processed using an Excel macro which extracted gender, age category and age graded performance score for all participants [34]. Age is provided as a 5-year cohort for adults except for 18-19 year olds. Age was converted to a continuous variable by assigning participants the mid-point for their cohort group. Only adult participants were included in the study. Gender is assigned by participants. Only participants selecting male or female were included in the study as age graded performance scores are not provided by parkrun for other gender options. The Scottish Index of Multiple Deprivation (simd) 2020 was determined for the data zone within which each parkrun is located [35]. Scotland is separated into 6,976 data zones which are ranked from the most deprived to the least deprived, based upon income, employment, education, health, access to services, crime and housing [35]. Event size was calculated from the mean attendance at each event for the year prior to the pandemic.

### Data analysis

The data were analysed using R x64 4.1.1 [36]. A generalised linear mixed model (GLMM) of event attendance in Scotland in the year prior to lockdown and the first year after return from lockdown was generated using the lme4 function [37]. Prior to modelling event attendances were transformed to natural logarithms. This not only made the data fit a Gaussian distribution but also allowed direct comparison between the rate of change in attendance across events of different sizes. All continuous explanatory variables (date, mean event size and simd ranking) were scaled to have a mean of zero and a standard deviation of one. In addition, a binary variable denoting whether the event was pre- or post-lockdown was included. Event location was included as a random effect. Minimum Akaike Information Criterion was used to select the optimal model [37].

GLMMs were generated for gender, age and performance of participants for the participant dataset. Event location was again used as random effect. A binomial error distribution was used for the model of gender. The simd of the area within which event is located and the mean attendance for the year prior to lockdown were included as predictor variables after scaling to have a mean of 0 and a standard deviation of 1.

## Results

### The impact of the COVID-19 lockdown on attendance at Scottish parkrun events

Prior to lockdown attendances in Scotland had been increasing exponentially (Figure 1) but a comparison of mean total attendance for the last year of parkrun in Scotland prior to the pandemic (7,649, 95% CI 7,275 to 8.024) and the first-year post-lockdown (6,612, 95% CI 6,260 to 6,963) reveals a more than 13% decline. A model of the attendance at individual events revealed a significant interaction between lockdown and event size revealing that declines in attendance were disproportionately larger at larger events (*β*=--0.039; p<0.001).

**Fig 1.**
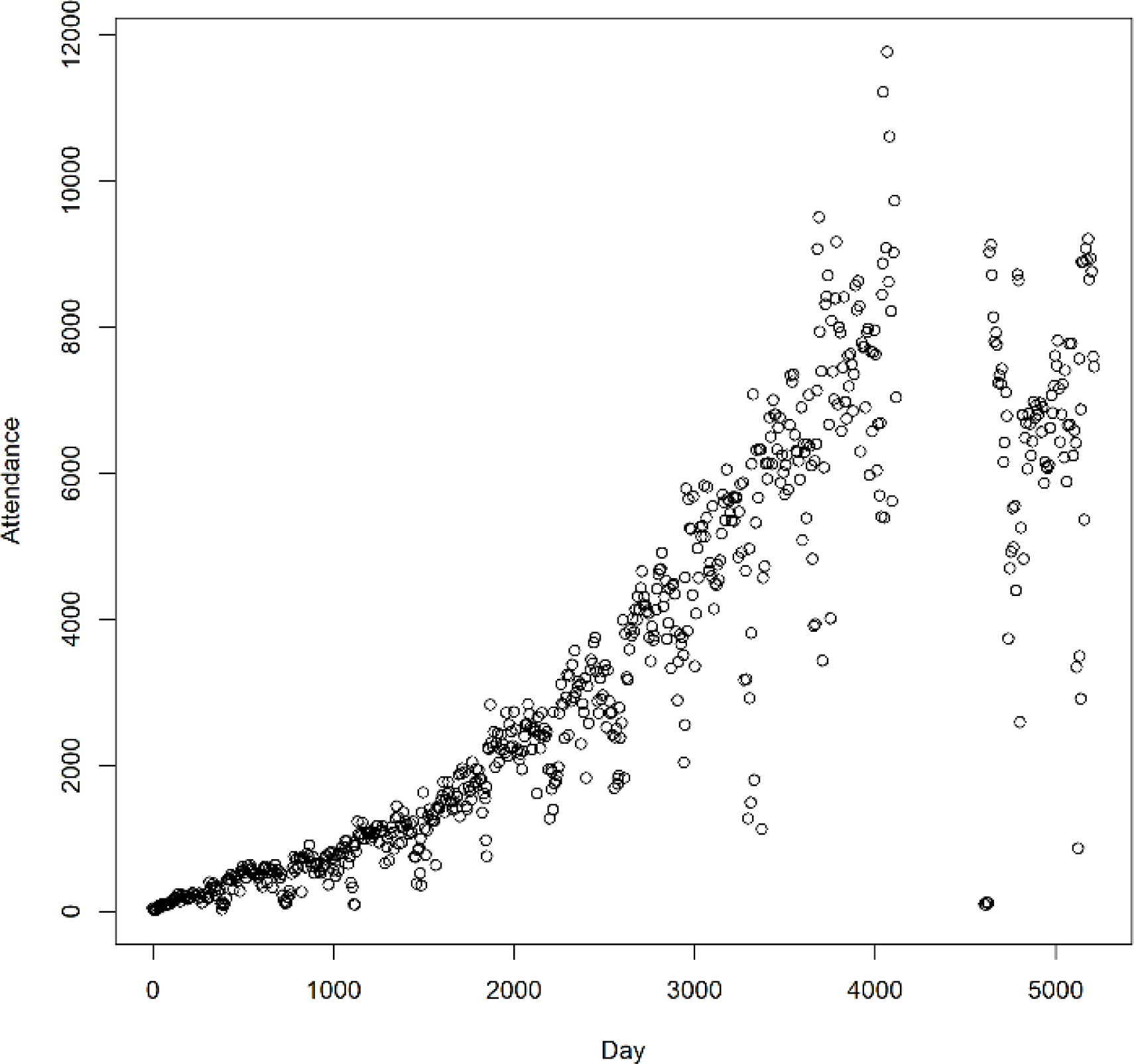
Total attendance on each parkrun day in Scotland from day 0, 6^th^ December 2008 until March 18^th^ 2023.

### The impact of COVID-19 lockdown on the gender ratio of participants at Scottish parkrun events

A total of 554,696 participations at parkrun events in Scotland a year either side of lockdown were included in the models consisting of 305,123 male participations and 249,573 female. Prior to lockdown the gender gap in participation at Scottish parkrun events had been narrowing [22]. A generalised linear model of gender ratio using a binomial error distribution and including event location as a random effect revealed a highly significant increase (*β*=0.088; p<0.001) in the gender ratio from 54.02% male (95% CI 53.85 to 54.19%) in the year prior to lockdown to 56.29% male (95% CI 56.09 to 56.49%) in the year after lockdown. An interaction term identified a significantly greater increase in gender ratio at larger events after lockdown (*β*=0.080; p=0.007). Separating event size into small and large based upon whether the scaled score for each event was positive (high) or negative (low) shows an increase in the gender ratio from 52.87% male (95% CI 52.63 to 53.10) before lockdown to 54.96% male (95% CI 54.70 to 55.23) after lockdown at small events and an increase in the gender ratio from 55.44% male (95% CI 55.18 to 55.70) before lockdown to 54.96% male (95% CI 57.67 to 58.27) after lockdown at large events.

### The impact of COVID-19 lockdown on the age of participants at Scottish parkrun events

There had been a trend towards an increase in the age of participants prior to lockdown in Scotland and a narrowing age gap between the genders [22]. A GLMM (Table 1) of the age of participants identified an increase in age of participants from 44.94 (95% CI 44.90 to 44.98), in the year before lockdown to 46.58 (95% CI 46.53 to 46.64) in the year after lockdown. A previous study reported an age gap between the sexes with male participants being on average older than female participants [22]. The model found the age gap between the genders has widened after lockdown (males: before lockdown 46.08 (95% CI 46.02 to 46.14), after lockdown 47.75 (95% CI 47.68 to 47.82); females: before lockdown 43.61 (95% CI 46.54 to 46.67), after lockdown 45.08 (95% CI 45.01 to 45.16). Larger events had a younger demographic, and this was more pronounced in female participants. More deprived areas had an older demographic than less deprived areas prior to lockdown although this association reversed after lockdown in female participants (Table 2). There was also a disproportionate loss of female participants in more deprived areas and male participants in less deprived areas after lockdown (χ^2^=6.26; p=0.012, Table 2).

**Table 1.** A general linear mixed model of the age of participants at Scottish parkrun events for the year before and the year after lockdown. Event location was included as a random effect. All continuous explanatory variables were scaled. Event size was the mean attendance at each event over the last year prior to lockdown.

| Parameter | Estimate | SE | P value |
| --- | --- | --- | --- |
| Intercept | 43.58 | 0.279 | <0.001 |
| Gender (Male) | 2.563 | 0.045 | <0.001 |
| Lockdown (Post) | 1.442 | 0.050 | <0.001 |
| Event size | -1.126 | 0.288 | <0.001 |
| Simd | 0.437 | 0.259 | 0.010 |
| Gender (Male)*Event size | 0.443 | 0.035 | <0.001 |
| Lockdown (Post)* Simd | -0.173 | 0.051 | <0.001 |
| Lockdown (Post)*Gender (Male) | 0.199 | 0.068 | 0.003 |
| Gender (Male)*Simd | -0.002 | 0.046 | 0.962 |
| Lockdown (Post)*Gender (Male)*Simd | 0.286 | 0.068 | <0.001 |

**Table 2.** The mean age and gender of participants at parkrun in Scotland in the year before and after lockdown in low and high deprivation areas. Simd was separated into high and low deprivation based upon whether the scaled score for each event was positive (low) or negative (high).

| Lockdown | Simd | Gender | N | Mean | 95% CI |
| --- | --- | --- | --- | --- | --- |
| Pre | high | Female | 70 578 | 43.46 | 43.37 to 43.55 |
| Post | high | Female | 52 041 | 45.16 | 45.05 to 45.27 |
| Pre | high | Male | 84 190 | 45.90 | 45.82 to 45.98 |
| Post | high | Male | 66 929 | 47.54 | 47.44 to 47.64 |
| Pre | low | Female | 73 529 | 43.74 | 43.65 to 43.82 |
| Post | low | Female | 53 425 | 45.00 | 44.89 to 45.11 |
| Pre | low | Male | 85 104 | 46.26 | 46.17 to 46.34 |
| Post | low | Male | 68 900 | 47.95 | 47.85 to 48.04 |

### The Impact of COVID-19 lockdown on the Performance of Participants at Scottish parkrun Events

There has been a trend towards a reduction in the performance of participants prior to lockdown in Scotland [22] as parkrun has become more inclusive to the less active. After lockdown that trend reversed with a GLMM (Table 3) identifying a highly significant increase in performance from 55.02% (95% CI 54.96-55.06%) pre-lockdown to 55.93% 95% CI 55.88 to 55.97%) after lockdown. The model identified that performance was positively correlated with age, being male and event size. Significant interaction terms revealed a narrowing gender gap in performance with increasing age, event size and levels of deprivation, together with a narrowing of the gender gap in performance after lockdown (males: before lockdown 56.90%; 95% CI 56.85-56.95, after lockdown 57.49; 95% CI 57.43 to 57.54; females: before lockdown 52.82; 95% CI 52.76 to 52.87, after lockdown 53.92; 95% CI 53.85 to 53.98). The model also revealed an increase in the performance gap between high and low deprivation areas after lockdown (Table 4).

**Table 3.** A general linear mixed model of the age graded performance score of participants at Scottish parkrun events for the year before and the year after lockdown. Event location was included as a random effect. All continuous explanatory variables were scaled. Event size was the mean attendance at each event over the last year prior to lockdown.

| Parameter | Estimate | SE | P value |
| --- | --- | --- | --- |
| Intercept | 53.21 | 0.319 | <0.001 |
| Age | 2.914 | 0.027 | <0.001 |
| Gender (Male) | 3.485 | 0.036 | <0.001 |
| Lockdown (Post) | 0.798 | 0.041 | <0.001 |
| Event size | 1.014 | 0.326 | 0.003 |
| Simd | -0.188 | 0.302 | 0.536 |
| Gender (Male)*Age | -1.163 | 0.037 | <0.001 |
| Gender (Male)*Event size | -0.484 | 0.028 | <0.001 |
| Lockdown (Post)*Gender (Male) | -0.404 | 0.054 | <0.001 |
| Lockdown (Post)*Age | 0.255 | 0.041 | <0.001 |
| Gender (Male)*Simd | 0.156 | 0.028 | <0.001 |
| Lockdown (Post)*Event size | 0.149 | 0.031 | <0.001 |
| Lockdown (Post)*Simd | -0.117 | 0.028 | <0.001 |
| Simd*Event size | 0.312 | 0.289 | 0.286 |
| Lockdown (Post)*Gender (Male)*Age | -0.420 | 0.054 | <0.001 |
| Lockdown (Post)* Simd*Event size | 0.127 | 0.026 | <0.001 |

**Table 4.**
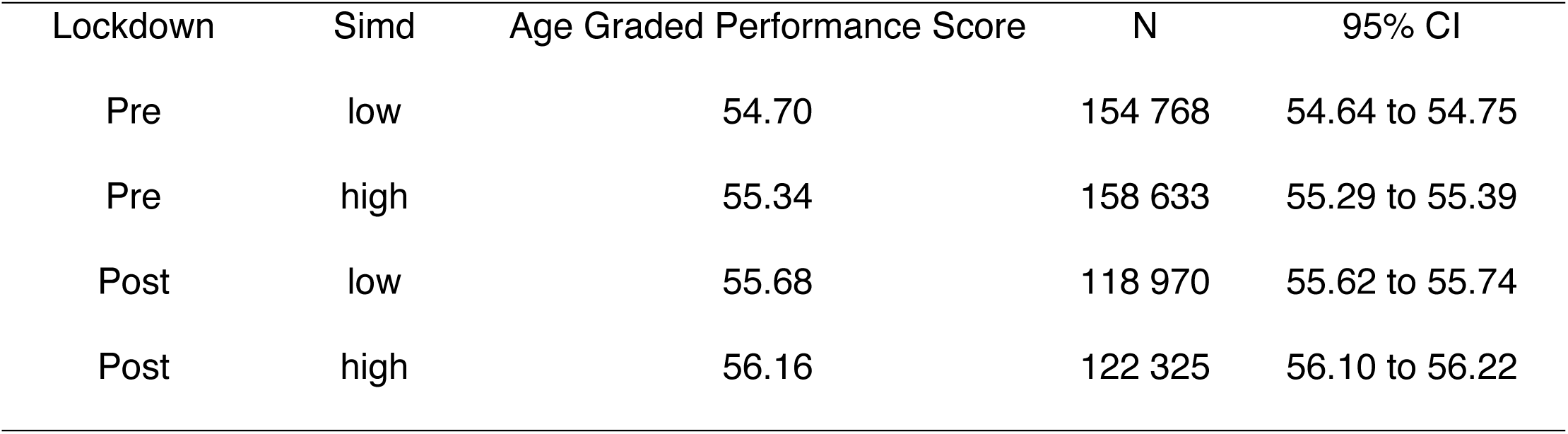
The mean age graded performance score of participants at parkrun events in Scotland in the year before and after lockdown. Events were separated into low and high simd based upon whether the scaled score for each event was positive (high) or negative (low).

| Lockdown | Simd | Age Graded Performance Score | N | 95% CI |
| --- | --- | --- | --- | --- |
| Pre | low | 54.70 | 154 768 | 54.64 to 54.75 |
| Pre | high | 55.34 | 158 633 | 55.29 to 55.39 |
| Post | low | 55.68 | 118 970 | 55.62 to 55.74 |
| Post | high | 56.16 | 122 325 | 56.10 to 56.22 |

## Discussion

The findings are consistent with previous studies that have identified that COVID-19 pandemic lockdowns reduced levels of physical activity particularly for the young, the less fit and those in most deprived areas [4–9,29]. The current study has identified that these patterns have continued into the post-lockdown period in a mass participation event in Scotland. The study has also identified some new correlates of activity and new patterns of association in existing correlates.

Overall attendances are well down on where they would have been had they continued to increase at pre-pandemic rates. The models identify several changes in correlates with participation levels after lockdown which help identify the changing demographics of parkrun attendees. A previous study identified that among parkrun participants from before the pandemic that the activity levels had particularly declined in younger adults, the least active and those in more deprived areas during lockdown [29].The current study reveals the concerning finding that these impacts also extend into the post-lockdown period. Furthermore, additional impacts were identified with a substantial shift in the gender ratio with a higher rate of loss of female participants. These means that several existing health inequalities have now only been exacerbated by lockdown but also that these impacts have extended into the post-lockdown era.

A number of important correlates of levels of participation were identified that help shed light on why some of these health inequalities have increased. The widening age gap between the genders suggests that there has been a bias towards the loss of females amongst older participants. The narrowing performance gap between the genders suggests that it is the least fit female participants that have stopped attending. The widening gender age gap at larger events suggests that there has also been a bias in the loss of older females from larger events. This seems to be particularly the case in the least deprived parts of Scotland. This could be related to older, less fit women avoiding more crowded parkruns to reduce their risk of contracting COVID-19. Both age and fitness are associated with the increasing risks of contracting COVID-19 [38,39] so it appears that older, less fit females could be more risk averse and are avoiding busier events. This is consistent with a study in Kenya that found that women in higher social classes were the most likely to reduce their levels of physical activity during lockdown [14]. The increasing gender gap in age at events in the least derived areas could be related to better awareness of the risks of contacting covid. The finding that smaller events had the greatest increase in age also suggests that older participants have altered their attendance patterns to avoid large crowds. This suggests that parkrun and other similar event organisers should continue to provide additional events in areas where attendances at existing events are high.

The study revealed a bias in age of participants with a younger demographic at larger parkruns and those in more deprived areas. The lack of places to exercise in more deprived areas might have pushed younger adults out of the exercise habit during lockdown [6–8]. These findings suggest it is increasingly important for governments to put in place measures to encourage younger adults to participate in more physical activity post-lockdown [5]. The same can be applied to those from more deprived areas and the less fit. parkrun is an extremely effective and relatively cheap mechanism of improving public health and well-being so government interventions to help parkrun make up for the time lost to the pandemic and reverse its impacts are now required. Studies have shown that the health benefits of moderate and mild sport are particularly high and that there is likely to be great value gained from organisations such as parkrun in stimulating the less active to engage in more activity, so public funding to support and aid recovery after lockdown is key to mitigate the impacts of the pandemic [40]. Furthermore, a study in China has shown that physical activity before the pandemic significantly improved the outcome of those infected with COVID-19 showing a community and population benefit to physical activity which makes the population more resilient to future waves of COVID-19 and other potential pandemic events [38]. Studies have also shown for older adults how important the social aspect of being part of a regular exercise group, this is something that parkrun excels at providing [15]

Studies have also found that those in more vulnerable groups were the most likely to reduce their physical activity levels and avoid green spaces during lockdown [39]. The findings of the current study are entirely consistent with this as it is the lowest performing parkrun participants from before lockdown who are the least likely to have returned, yet they are those most likely to benefit from participation. There needs to be approaches developed to encourage those who reduced their level of physical activity as a result of fear of catching COVID-19 back into engaging with more exercise. One key demographic affected by this appears to be older women exacerbating another existing health inequality. More studies are needed into the factors driving these demographic changes to better understand and put in place measures to mitigate the impacts of lockdown of levels of physical activity.

## Conclusion

This large-scale study reveals that the impacts of the pandemic on levels of physical activity have continued into the post-lockdown era. These impacts are increasing health inequalities affecting women, younger adults, the least fit and are having different impacts based upon the level of social deprivation within an area. The study also shows how valuable the parkrun results database can be for uncovering the demographics of engagement with a mass participation event.

With respect to parkrun the pandemic has had a number of negative impacts in Scotland. It has reduced attendances, it has increased the gender ratio towards males, it has increased the mean age of participants and their performance levels. This suggests that the pandemic has undone some of the important work that parkrun had done to increase inclusivity. Prior to the pandemic the gender ratio of participants was moving toward equality and the performance levels dropping year on year as parkrun became more inclusive and successfully removed barriers to women and the least fit taking part in exercise. The pandemic has reversed these trends. As parkrun is a very cheap and effective mechanism of increasing fitness and reducing burdens of healthcare systems government investment into parkrun is needed to help them overcome the negative impacts of the pandemic and help communities become more resilient to future pandemics [41].

## Ethics approval

The ethics approval was obtained from the ethics review board of the University of Stirling (Reference No. EC 2023 14083 9598). There were no active participants in the study. This was an analysis of aggregated publicly available secondary datasets.

## Availability of data and materials

The datasets used in the study are available in the University of Stirling DATAStorre.

## Data Availability

The data will be made available in the Stirling DataSTORRE online repository

## Acknowledgements

The author acknowledges the use of data owned by parkrun Global. The data have been accessed as a permitted act for independent non-commercial research purposes through fair dealing legislation allowing access to publicly available databases. Only a tiny proportion of the parkrun results database was accessed (data from just 44 of more than 2000 events).

